# The role of Late Presenters on HIV-1 transmission clusters in Europe

**DOI:** 10.1101/2023.03.14.23287098

**Authors:** Mafalda N. S. Miranda, Marta Pingarilho, Victor Pimentel, Perpétua Gomes, Maria do Rosário O. Martins, Sofia G. Seabra, Rolf Kaiser, Michael Böhm, Carole Seguin-Devaux, Roger Paredes, Marina Bobkova, Maurizio Zazzi, Francesca Incardona, Ana B. Abecasis

**Affiliations:** Global Health and Tropical Medicine (GHTM), Institute of Hygiene and Tropical Medicine, New University of Lisbon (IHMT/UNL), 1349-008 Lisbon, Portugal; Laboratório de Biologia Molecular (LMCBM, SPC, CHLO-HEM), Lisbon, Portugal; Centro de Investigação Interdisciplinar Egas Moniz (CiiEM), Instituto Universitário Egas Moniz, Costa da Caparica, Portugal; Institute of Virology, Faculty of Medicine and University Hospital of Cologne, University of Cologne, Germany; DZIF Deutsches Zentrum für Infektionsforschung; Laboratory of Retrovirology, Department of Infection and Immunity, Luxembourg Institute of Health, Esch-sur-Alzette, Luxembourg; Infectious Diseases Department and IrsiCaixa AIDS Research Institute, Hospital Universitari Germans Trias i Pujol, Badalona, Spain; Gamaleya National Research Center of Epidemiology and Microbiology, Moscow, Russia; Department of Medical Biotechnologies, University of Siena, Siena, Italy; IPRO—InformaPRO S.r.l., Rome, Italy; EuResist Network, Rome, Italy

**Keywords:** HIV-1 infection, Late presenters, Non-Late Presenters, Transmission clusters

## Abstract

**Background:** Investigating the role of late presenters (LP) on HIV-1 transmission is important, as they can contribute to the onward spread of HIV-1 virus in the long period before diagnosis, when they are not aware of their HIV status

**Objective:** To describe the clinical and socio-demographic characteristics of HIV-1 infected individuals followed in Europe, to characterize patients in clusters and to compare transmission clusters (TC) in LP vs non-late presenters (NLP) populations.

**Methods:** Clinical, socio-demographic and genotypic information from 38531 HIV-1 infected patients was collected from the EuResist Integrated Database (EIDB) between 1981 and 2019. Sequences were aligned using VIRULIGN. Maximum likelihood (ML) phylogenies were constructed using FastTree. Putative transmission clusters were identified using ClusterPicker v1.332. Statistical analyses were performed using R.

**Results:** 32652 (84·7%) sequences were from subtype B, 3603 (9·4%) were from subtype G, and 2276 (5·9%) were from subtype A. The median age was 33 (IQR: 26·0-41·0) years old and 75·5% of patients were males. The main transmission route was through homosexual (MSM) contact (36·9%) and 86·4% were originated from Western Europe. Most patients were native (84·2%), 59·6% had a chronic infection, and 73·4% had acquired drug resistance (ADR). CD4 count and viral load at diagnosis (log10) presented a median of 341 cells/mm3, and of log10 4·3 copies/mL, respectively. 51·4% of patients were classified as LP and 21·6% patients were inside TCs. Most patients from subtype B (85·6%) were in clusters, compared to subtypes A (5·2%) and G (9·2%). Phylogenetic analyses showed consistent clustering of MSM individuals. In subtype A, patients in TCs were more frequently MSM patients and with a recent infection. For subtype B, patients in TCs were more frequently those with older age (≥ 56), MSM transmission route, originating from Western Europe, migrants, and with a recent infection. For subtype G, patients in TC were more frequently patients with recent infection and migrants. When analysing cluster size, we found that NLP more frequently belonged to large clusters (>8 patients) when compared to LP.

**Conclusion:** While late presentation is still a threat to HIV-1 transmission, LP individuals are more present either outside or in small clusters, indicating a limited role of late presentation to HIV-1 transmission.

## Introduction

At the end of 2020, there were 37·7 million people living with HIV (1) and it is known that in HIV epidemics, certain risk groups contribute to the spread of HIV disproportionately more than others. This can be due to specific demographic, clinical or behavioral factors or to factors related to the infecting strain of the virus (2,3). On the one hand, the literature suggests that a recent infection, without diagnosis, could be associated with transmission and spread of HIV-1 (4). On the other hand, late presentation to diagnosis has been increasing over the years and in Europe, late presenters (LP) account for around 50% of HIV new diagnosis (5). Late presenters (LP), based on a definition consensus, are HIV-1 infected individuals defined by a baseline CD4 count lower than 350 cells/mm3 or with an AIDS-defining event, regardless of CD4 cell count (6). Late presentation is associated with high morbidity and mortality, at an individual level, and increased health costs (7). Besides those consequences, LP can also contribute to the onward spread of HIV-1 virus at the population level, as these individuals are not aware of their HIV status and could also spread the virus without knowing their infection status (8).

The use of powerful tools as phylogenetic trees and transmission clusters (TC) are essential to understand the dynamics of viral transmission and to identify groups of individuals connected to each other (2,9).

In this study, we aim to describe the clinical and socio-demographic characteristics of HIV-1 infected individuals followed in Europe according to subtype and to understand the determinants associated with clustering on each of the more prevalent subtypes. The analysis of transmission clusters for the most prevalent HIV-1 subtypes, B, A, and G, and comparison of the patterns of transmission clusters in late presenters (LP) vs non-late presenters (NLP) populations were included in this study.

## Methods

### Study Group

Clinical, socio-demographic and genomic information from 38531 HIV-1 infected patients from the EuResist Integrated Database (EIDB) between 1981 and 2019 were included in this study. The EuResist integrated database (EIDB) is one of the largest existing datasets which integrate clinical, socio-demographic and viral genotypic information from HIV-1 patients. It integrates longitudinal, periodically updated data mainly from Italy (ARCA database), Germany (AREVIR database) Spain (CoRIS and IRSICAIXA), Sweden, Belgium, Portugal, and Luxembourg databases (10).

In this study, information from the ARCA, AREVIR, Luxembourg, IRSICAIXA, Portugal and Russia databases were used.

### Institutional Review Board Statement

All procedures performed in this study were in accordance with the ethical standards of the ethical committee of the Instituto de Higiene e Medicina Tropical, Universidade Nova de Lisboa and with the 1964 Helsinki declaration. The database enrolled anonymized patients’ information, including demographic, clinical and genomic data from patients from the EuResist Integrated Database (Date of approval: 15 January 2021).

### Drug Resistance Analysis and Subtyping

HIV *pol* sequences were derived from existing routine clinical genotypic resistance tests (Sanger method). The size of reverse transcriptase (RT) and protease (PR) fragments used for this analysis was between 500 and 1000 nucleotides. Only the first HIV genomic sequence per patient was analyzed. Transmitted Drug Resistance (TDR) was defined as the presence of one or more surveillance drug resistance mutations in a sequence, according to the WHO 2009 surveillance list (11). The sequences were submitted to the Calibrated Population Resistance tool version 8.0. Clinical resistance to ARV drugs was calculated through the Standford HIVdb version 9.0.

HIV-1 subtyping was performed using the consensus of the result obtained based on three different subtyping tools: REGA HIV Subtyping Tool version 3.46 (https://www.genomedetective.com/app/typingtool/hiv), COMET: adaptive context-based modeling for HIV-1 (https://comet.lih.lu) and SCUEAL (“http://classic.datamonkey.org/dataupload_scueal.php“).

### Transmission cluster (TC) identification

For the analysis of transmission clusters and construction of phylogenetic trees, the database was divided in three separate datasets, subtype B, A, and G. Control sequences were retrieved from the Los Alamos database and all HIV-1 *pol* sequences from subtypes B, A, and G from Europe, South America and Africa were included (http://www.hiv.lanl.gov) (12). We used as the outgroup reference three subtype B and C references retrieved from the Los Alamos database. For each subtype, sequences were aligned against the control sequences dataset using VIRULIGN (13). The HIV-1 K03455.1 (HXB2) pol nucleotide sequence (nt) was used as reference for codon corrected alignment. The dataset was then manually edited to exclude sequences with low quality, duplicates and clones using MEGA7 software. The final datasets of subtypes B, A, and G consisted of 62543, 10122, and 5547 sequences, respectively, with a length of 948. Maximum likelihood (ML) phylogenies were constructed in FastTree with the generalized time reversible model. Statistical support of clades was assessed using the Shimodaira-Hasegawa-like test (SH-test). Putative transmission clusters were identified using ClusterPicker v1.332 (14) and defined a threshold that included a genetic distance of 0.030 and a branch support ≥0·90 aLRT. For analyses of cluster size, we defined clusters with 8 patients or more as large clusters and cluster with less than 8 patients as small clusters. The visual configuration of the phylogenetic trees was possible through the iTOL programme.

### Study Variables

New variables were created according to:

- Migration Status- Based on Country of Origin and Country of Follow-up (if country of origin and country of follow-up is the same, then patient was classified as native; otherwise as migrant)
- Age at Resistance Test - Based on the difference between Year of Birth and Date of the first drug resistance test;
- Region of Origin- Based on Country of Origin;
- Treatment Status at date of first Drug Resistance Test – based on the difference between sample collection date for first drug resistance test and date of start of first therapy: ART-naïve→ patients who had a sample collection date for first drug resistance test before the date of start of first therapy ART-experienced→ patients who had a sample collection date for first drug resistance test after the date of start of first therapy
- Recentness of infection - Based on ambiguity rate of genomic sequences. We defined Chronic Infection if the ambiguity rate was higher than 0·45% otherwise Recent infection was defined, as previously described (15).
- LP vs NLP at HIV diagnosis- Based on CD4 count, LP were defined as patients with a baseline CD4 count =< 350 cells/mm3 and NLP were defined as patients with baseline CD4 count > 350 cells/mm3 (6).

### Statistical analysis

The proportion and median (interquartile range, IQR) were calculated for every categorical and continuous variable, respectively. The treatment status variable was compared with the categorical variables with Chi-square test, and continuous variables with Mann-Whitney U test. Logistic regression was used to analyze the association between demographic, clinical factors, clustering status and the subtypes. First, we presented the logistic regression with the unadjusted odds ratios (uOR) and confidence intervals at 95% (95% CI), then we included only the variables with a p-value <0·05 in the final model. The final model was adjusted for sex, this variable was forced into the model regardless of its significance. Data was analyzed using RStudio (Version 1.2.5033).

## Results

### Characteristics of European Population

Among the 38531 HIV-1 infected patients from the EIDB included in the analysis, 32652 (84·7%) were from subtype B, 3603 (9·4%) were from subtype G and 2276 (5·9%) were from subtype A. The median age at resistance test was 40·0 (34·0-47·0) years old and 75·5% of HIV-1 infected patients were males. The main transmission route was through homosexual (men who have sex with men-MSM) contact (36·9%). For subtype B, the MSM route (40·7%) was also the most prevalent route whereas was the heterosexual route was the predominant route for subtype A and G (43·3% and 49·9%, respectively) (Data not shown). 86·4% of patients were originated from Western Europe and according to subtypes, South America was the most prevalent region of origin for subtype B, whereas subtype A-infected patients were mainly from Eastern Europe region and for G it was Africa region the most prevalent. Most patients included in this study were native (84·2%) and according to subtype, in subtype B natives were more prevalent (94·8%) and in subtype A and G migrants were more prevalent (13·8% and 16·6%). Based on the ambiguity rate of the first genomic sequence, most patients were classified as presenting a chronic infection (59·6%). Most patients were ART-experienced (59%) and 73·4% had acquired drug resistance (ADR). CD4 count at diagnosis and viral load at diagnosis (log10) presented a median of 341 cells/mm3 (IQR 170-540) and log10 4·3 copies/mL (IQR 3·4-5·0), respectively. 21·6% of patients were represented within transmission clusters and 51·4% of patients were classified as LP (CD4<350 cells/mm3). Most patients from subtype B (85·6%) were located in clusters in contrast to subtypes A (5·2%) and G (9·2%) (Table S1).

### Dynamics of subtype A HIV-1 epidemic in Europe

Based on the sequences from our database and the control sequences retrieved, we could observe that the majority of the subtype A population had its origin in Africa, and the major route of transmission was heterosexual. There were some individuals with IDU transmission. The phylogenetic analyses indicates that most EuResist patients cluster in two different parts of the tree, indicated with arrows A and B in figure 1, suggesting two parallel epidemics of subtype A in Europe. The first cluster was related to patients originating from Africa and Eastern Europe through heterosexual and IDU transmission (cluster A) and the other was linked to patients originating from Western Europe with MSM transmission (cluster B). LP individuals are mostly concentrated in cluster A, where the majority of individuals are also migrants (Figure 1).

**Figure 1.**
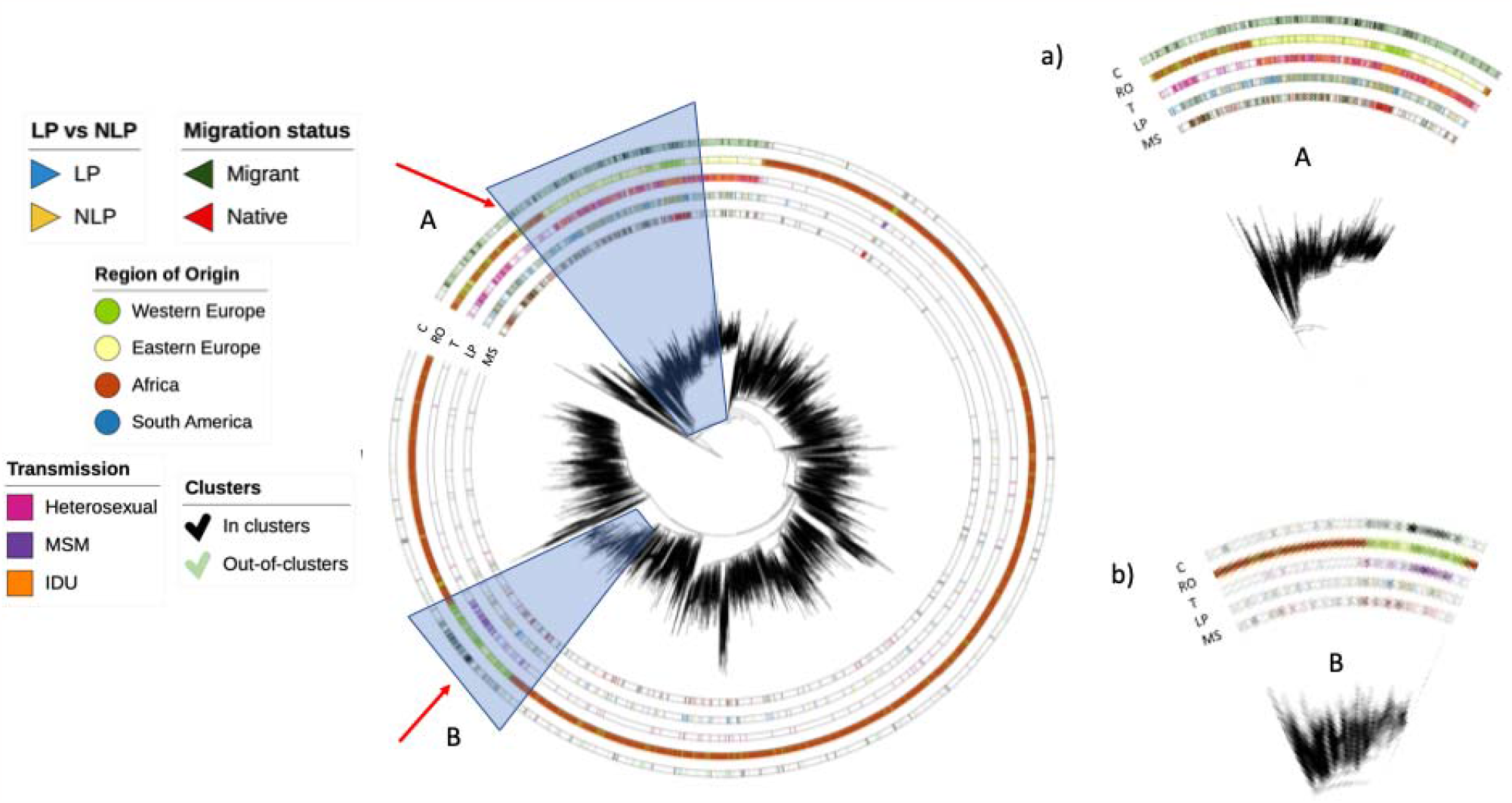
Phylogenetic tree for Subtype A. This image shows a visual phylogenetic tree of the subtype A population. The region A, figure 1.a), shows a clustering of individuals originating from Africa and Eastern Europe with a heterosexual and IDU transmission, the region B, figure 1.b), shows a clustering of individuals originating from Western Europe and MSM transmission. C-Clusters; RO- Region of origin; T- Transmission; LP- Late presenters vs non-late presenters; MS- Migration status

### Dynamics of subtype B HIV-1 epidemic in Europe

Based on the sequences from our database and the control sequences retrieved, we could observe that most subtype B patients originated from Western Europe, are native and MSM. Individuals with IDU transmission originating from Western Europe dominate one cluster of the tree (indicated with an arrow). LP and NLP individuals are distributed evenly in the tree. Based on the configuration of the phylogenetic tree and apart from the cluster dominated by IDUs, there seems to be no major compartmentalization patterns in the subtype B epidemic in Europe (Figure 2).

**Figura 2.**
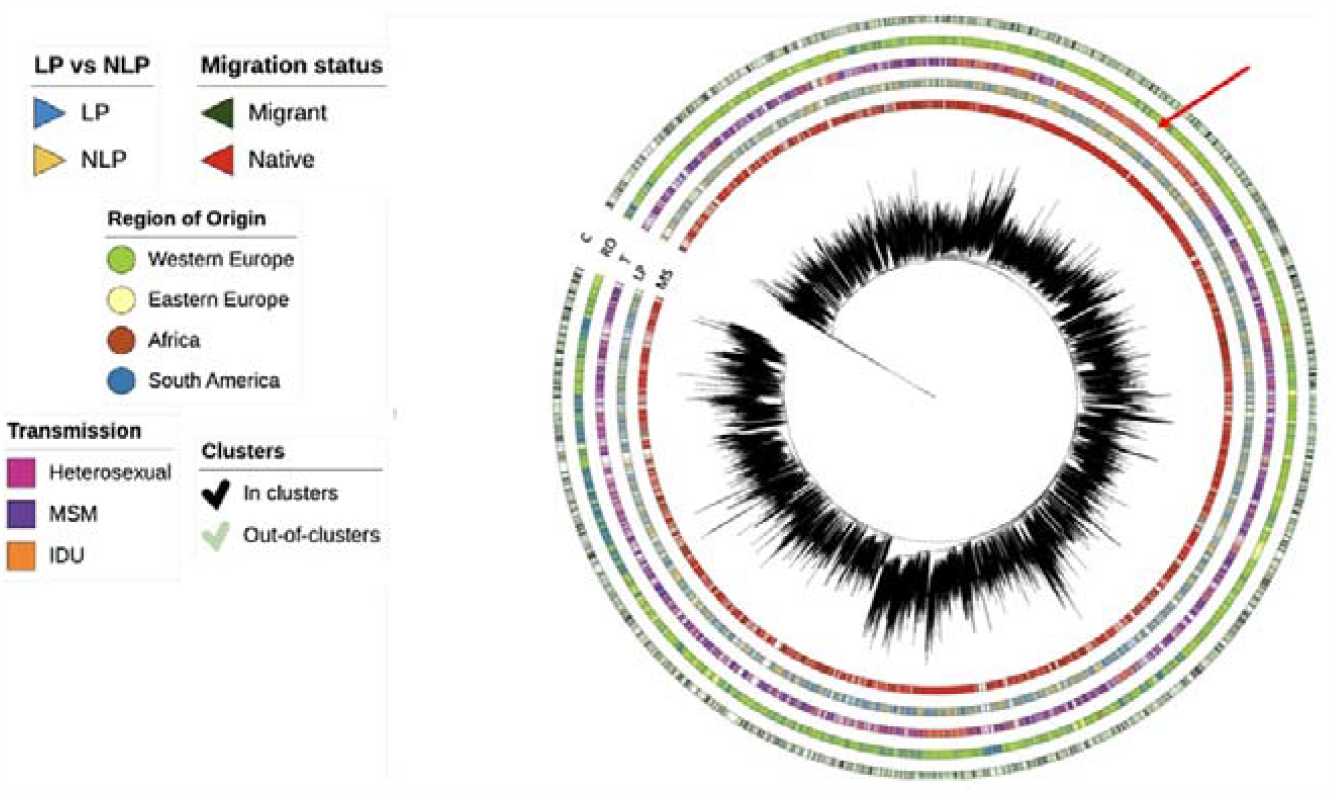
Phylogenetic tree for Subtype B. This image shows a visual phylogenetic tree of the subtype B population. The region highlighted with an arrow shows a clustering of individuals with an IDU transmission and originating from Western Europe. C- Clusters; RO- Region of origin; T- Transmission; LP- Late presenters vs non-late presenters; MS- Migration status

### Dynamics of subtype G HIV-1 epidemic in Europe

Based on the sequences from our database and the control sequences retrieved, we could observe two major regions of origin - Western Europe and Africa – compose the subtype G epidemic of HIV-1 in Europe. These are largely divided in two major clusters indicated with arrows A and B in the Figure 3. Most individuals are heterosexuals and LP dominate. The tree configuration indicates lack of compartmentalization of subtype G epidemic in Europe and suggests frequent importations of subtype G.

**Figure 3.**
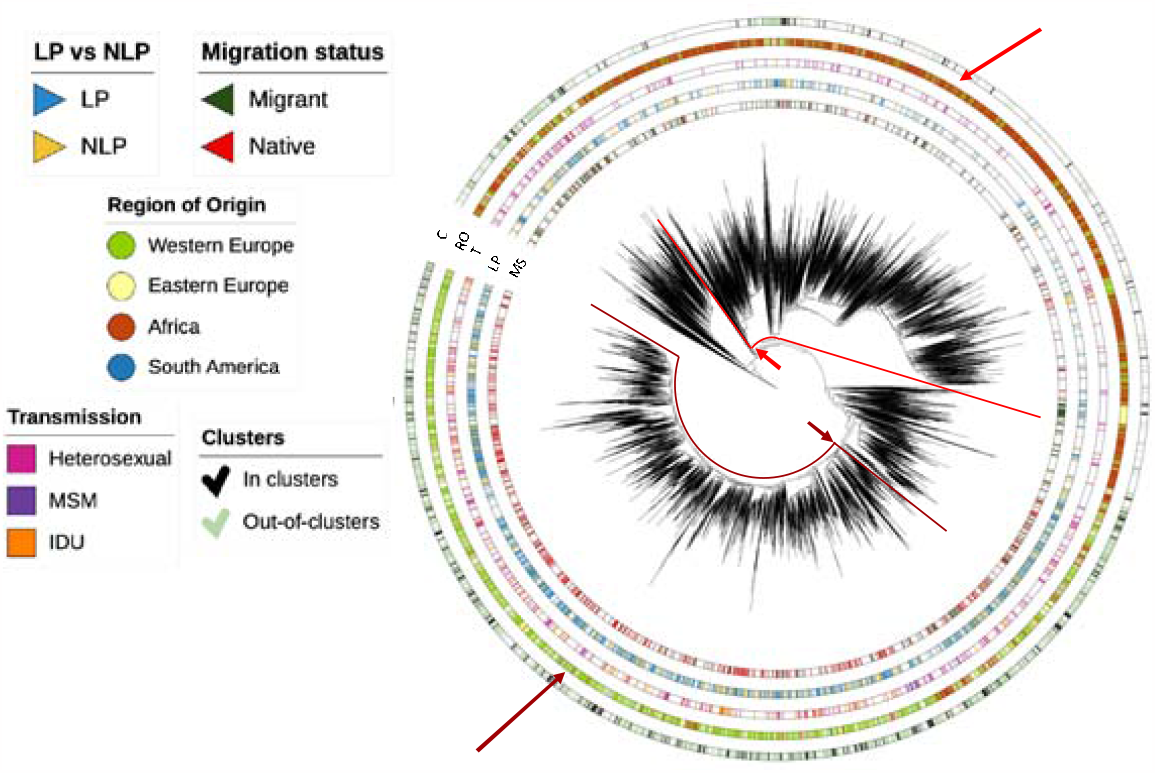
Phylogenetic tree for subtype G. This image shows a visual phylogenetic tree of the subtype G population. The regions highlighted with arrows shows a clustering of individuals originating from Western Europe and Africa. C- Clusters; RO- Region of origin; T- Transmission; LP- Late presenters vs non-late presenters; MS- Migration status

### Determinants associated with transmission clusters of HIV-1 in Europe for different subtypes

In the first unadjusted logistic regression model for Subtype A, the variables associated with a patient being in clusters from subtype A were male individuals (p<0·001), MSM (p<0·001) route of transmission, having a recent infection (p<0·001) and being NLP (p=0·004) (Table S2).

In the final logistic regression model for subtype A, we adjusted the model to the variable sex, and individuals with a MSM transmission route were more likely to be in clusters when compared to heterosexual route (OR:2·65, p=0·001). Patients with a recent infection were more likely to be in clusters when compared to individuals with a chronic infection (OR:2·70, p<0·001) (Table 1.).

**Table 1.** Determinants associated with belonging to a transmission cluster according to Subtype A, B and G.

|  |  | Subtype A |  | Subtype B |  | Subtype G |  |
| --- | --- | --- | --- | --- | --- | --- | --- |
| In clusters/Out-of-clusters |  | Final Model (Stepwise) |  | Final Model (Stepwise) |  | Final Model (Stepwise) |  |
|  |  | aOR (95% CI) | p-value | aOR (95% CI) | p-value | aOR (95% CI) | p-value |
| Sex | Female | Ref | Ref | Ref | Ref | Ref | Ref |
|  | Male | 1.44 (0.95-2.18) | 0.083 | 1.18 (1.01-1.38) | **0.037** | 0.80 (0.56-1.16) | 0.237 |
| Age at resistance test | <18 |  |  | 1.32 (0.70-2.50) | 0.391 |  |  |
|  | 19-30 |  |  | 1.49 (1.20-1.86) | **<0.001** |  |  |
|  | 31-55 |  |  | 1.05 (0.87-1.28) | 0.617 |  |  |
|  | >56 |  |  | Ref | Ref |  |  |
| Transmission Route | Heterosexual | Ref | Ref | Ref | Ref |  |  |
|  | MSM | 2.65 (1.50-4.69) | **0.001** | 1.74 (1.52-2.00) | **<0.001** |  |  |
|  | IDU | 0.79 (0.50-1.25) | 0.308 | 0.58 (0.49-0.68) | **<0.001** |  |  |
|  | Other | 0.91 (0.51-1.61) | 0.747 | 0.73 (0.53-1.01) | 0.057 |  |  |
| Region of Origin | Western Europe |  |  | Ref | Ref |  |  |
|  | Eastern Europe |  |  | 1.00 (0.69-1.46) | 0.997 |  |  |
|  | Africa |  |  | 0.75 (0.46-1.22) | 0.249 |  |  |
|  | South America |  |  | 0.30 (0.21-0.44) | **<0.001** |  |  |
|  | Other |  |  | 0.61 (0.39-0.95) | **0.028** |  |  |
| Migration Status | Migrant |  |  | Ref | Ref | Ref | Ref |
|  | Native |  |  | 0.60 (0.49-0.72) | **<0.001** | 1.55 (1.07-2.25) | **0.021** |
| Recentness of Infection | Chronic | Ref | Ref | Ref | Ref | Ref | Ref |
|  | Recent | 2.70 (1.88-3.88) | **<0.001** | 1.88 (1.70-2.07) | **<0.001** | 1.99 (1.37-2.87) | **<0.001** |

In the subtype B unadjusted logistic regression model, the variables associated with a patient being in clusters were male individuals (p<0·001), individuals with an age at resistance test between 19-30 years (p<0·001), route of transmission of MSM (p<0·001), patients originating from Eastern Europe (p<0·001), being migrant (p<0·001) and having a recent infection (p<0·001). Also, not having ADR (p<0·001), being NLP (p<0·001) and higher levels of viral load (p<0·001) were also associated with being in clusters from subtype B (Table S2).

The final logistic regression model was adjusted to sex and the determinants associated with a patient being in clusters from subtype B were males (OR:1·18, p=0·037), age at resistance testing, individuals with an age between 19-30 years were more likely to be within clusters (OR:1·49, p=0·002) when compared to older age (>56 years old). Individuals with a MSM transmission route were more likely to be in clusters when compared to heterosexual route (OR:1·74, p<0·001), while individuals with a IDU transmission route were less likely to be in clusters when compared to heterosexuals (OR:0·58, p<0·001). Patients originated from South America had a lower probability of being in clusters when compared to patients originated from Western Europe (OR:0·30, p<0·001). Native individuals were less likely to be in clusters when compared to migrants (OR:0·60, p<0·001) and individuals with a recent infection were more likely to be in clusters when compared to individuals with a chronic infection (OR:1·88, p<0·001) (Table 1.).

In the subtype G unadjusted logistic regression model, the variables associated with a patient being in clusters were female individuals (p=0·012), being native (p=0·015), having a recent infection (p<0·001), not having ADR (p<0·001) and being NLP (p=0·037) (Table S2).

The final logistic regression model was adjusted to sex and individuals from subtype G and in clusters were more likely to be native when compared to migrants (OR:1·55, p=0·021). Other factor associated with a patient being in clusters from subtype G was to have a recent infection when compared to those individuals with a chronic infection (OR: 1·99, p<0·001) (Table 1.).

### Transmission Clusters analysis

Of the 38531 patients, 8335 were in clusters (21·6%). The minimum clusters size was 2 and the maximum clusters size was 52 (data not shown).

The proportion of late presenters and non-late presenters in clusters was analyzed and LP were more in small clusters (95·7%) than NLP (92·4%). Also, LP within the small clusters were more in dual clusters (65·6%) (2 patients per clusters) (Figure 4.A).

**Figure 4.**
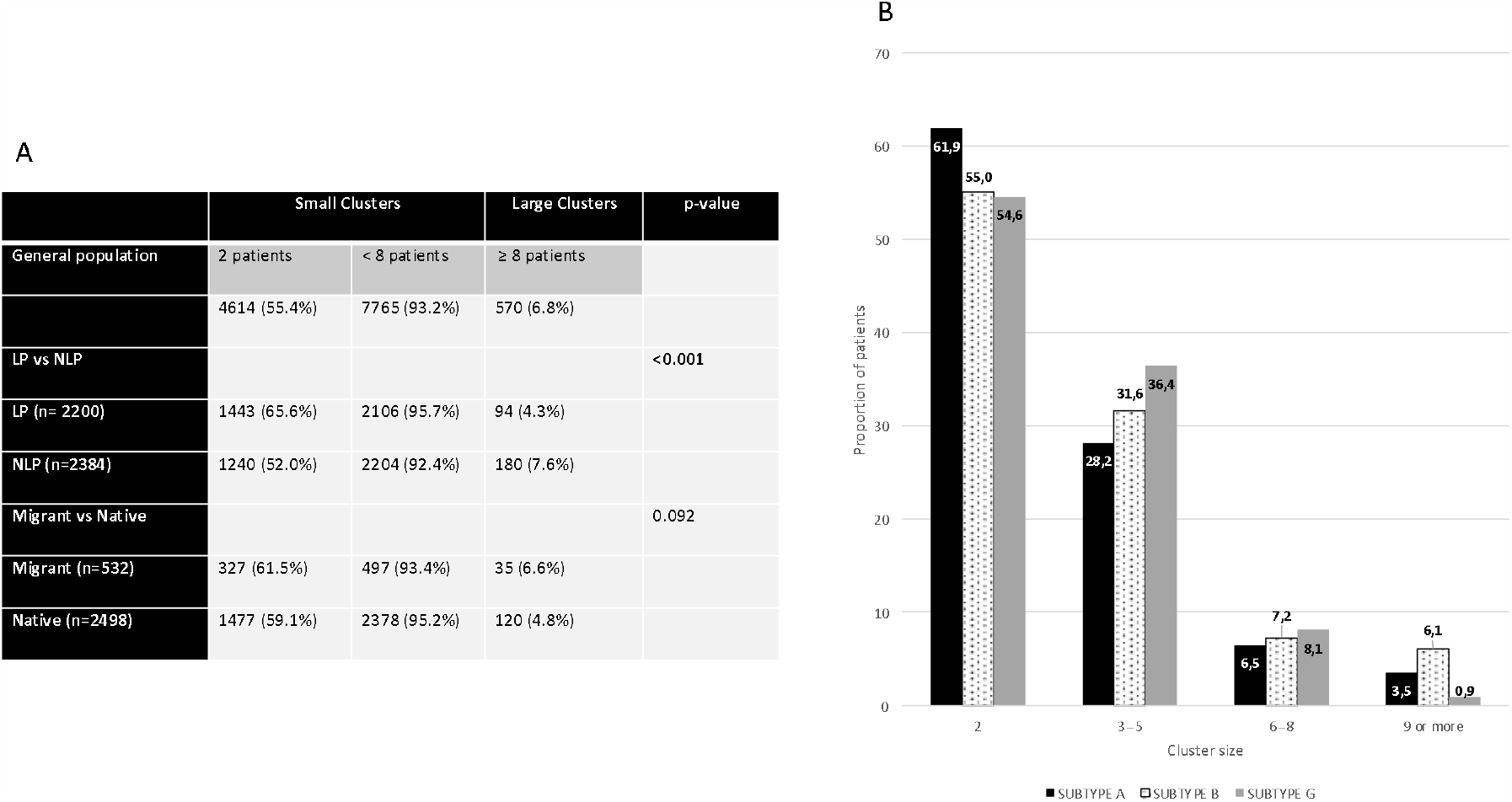
General clusters size characterization (A) and clusters size according to subtypes (B).

We also analyzed the proportion of migrants vs natives in clusters, and migrants were more in large clusters (6·6%) than natives (4·8%) (Figure 4.A).

According to subtypes, subtype A had a higher proportion of patients in dual clusters (61·9%), subtype B had a higher proportion of patients in clusters of 9 or more (6·1%) and subtype G has a higher proportion of patients in clusters between 3-5 and 6-8 clusters (36·4% and 8·1%, respectively) (Figure 4.B).

### Transmission Clusters in LP vs NLP

Here we studied the associated characteristics to LP vs NLP in clusters. Although, within the clinical and socio-demographic characteristics, there were specific characteristics of LP in clusters and NLP in clusters. LP were mainly out-of-clusters in all subtypes. In subtype A the variables associated with being LP vs NLP in clusters were age at resistance test (p=0·029) and recentness of infection (p=0·017), where LP individuals in clusters were older than 31yo (p=0·001; p=0·030), while NLP in clusters were males (p=0·002), with an age between 19-30yo (p=0·027) and a recent infection (p<0·001).

LP vs NLP with subtype B in clusters were associated with age at resistance testing (p<0·001), treatment status (p<0·001), transmission route (p=0·033), recentness of infection (p<0·001), viral load (p<0·001) and ADR (p=0·010). Where LP in clusters were older than 31yo (p<0·001; p<0·001), ART-experienced (p<0·001), originating from Eastern Europe (p<0·001), viral load higher than 5.1 copies/mL (p<0·001) and having ADR (p<0·001). While NLP in clusters were males (p<0·001), younger than 30yo (p<0·001;p<0·001), ART-naïves (p<0·001), with a MSM and IDU route of transmission (p<0·001 and p=0·047, respectively), from Western Europe (p<0·001), with a recent infection (p<0·001) and a viral load lower than 4.0 copies/mL (p<0·001). LP vs NLP with subtype G in clusters were associated with the variables treatment status (p=0·015), recentness of infection (p=0·001), and transmission route (p=0·035). LP in clusters were mainly females (p=0·002), ART-experienced (p<0·001), with a chronic infection (p<0·001) and an IDU transmission route (p=0·011). For this subtype, MSM route in clusters was 100% related to NLP, but without being statistically significant (Table 2.)

**Table 2.** Characteristics of LP and NLP in clusters according to subtypes.

|  | In clusters |  |  |  |
| --- | --- | --- | --- | --- |
|  | LP | NLP | p-value | p-value compared |
| **Subtype A** |  |  |  |  |
| **Sex** |  |  | 0.052 |  |
| Male (n=109) | 43 (39.4%) | 66 (60.6%) |  | **0.002** |
| Female (n=64) | 35 (54.7%) | 29 (45.3%) |  | 0.288 |
| **Age at resistance test** |  |  | **0.029** |  |
| <18 (n=4) | 1 (25%) | 3 (75%) |  | 0.157 |
| 19-30 (n=33) | 12 (36.4%) | 21 (63.6%) |  | **0.027** |
| 31-55 (n= 62) | 40 (64.5%) | 22 (35.5%) |  | **0.001** |
| >56 (n= 12) | 8 (66.7%) | 4 (33.3%) |  | **0.030** |
| **Recentness of infection** |  |  | **0.017** |  |
| Chronic (n=84) | 45 (53.6%) | 39 (46.4%) |  | 0.351 |
| Recent (n=95) | 34 (35.8%) | 61 (64.2%) |  | **<0.001** |
| **Subtype B** |  |  |  |  |
| **Sex** |  |  | 0.879 |  |
| Male (n=3421) | 1632 (47.7%) | 1789 (52.3%) |  | **<0.001** |
| Female (n=629) | 298 (47.4%) | 331 (52.6%) |  | 0.065 |
| **Age at resistance test** |  |  | **<0.001** |  |
| <18 (n=16) | 2 (12.5%) | 14 (87.5%) |  | **<0.001** |
| 19-30 (n=608) | 216 (35.5%) | 392 (64.5%) |  | **<0.001** |
| 31-55 (n=2307) | 1215 (52.7%) | 1092 (47.3%) |  | **<0.001** |
| >56 (n=245) | 154 (62.9%) | 91 (37.1%) |  | **<0.001** |
| **Treatment Status** |  |  | **<0.001** |  |
| ART-naive (n=2121) | 1014 (47.8%) | 1107 (52.2%) |  | **0.004** |
| ART-experienced (n=1284) | 707 (55.1%) | 577 (44.9%) |  | **<0.001** |
| **Transmission Route** |  |  | **0.033** |  |
| Heterosexual (n=712) | 373 (52.4%) | 339 (47.6%) |  | 0.070 |
| MSM (n=1626) | 760 (46.7%) | 866 (53.3%) |  | **<0.001** |
| IDU (n=427) | 199 (46.6%) | 228 (53.4%) |  | **0.047** |
| **Region of Origin** |  |  | 0.256 |  |
| Western Europe (n=2795) | 1332 (47.7%) | 1463 (52.3%) |  | **<0.001** |
| Eastern Europe (n=81) | 47 (58%) | 34 (42%) |  | **0.042** |
| Africa (n=53) | 26 (49.1%) | 27 (50.9%) |  | 0.853 |
| South America (n=88) | 46 (52.3%) | 42 (47.7%) |  | 0.542 |
| **Recentness of infection** |  |  | **<0.001** |  |
| Chronic (n=1942) | 1126 (58%) | 816 (42%) |  | **<0.001** |
| Recent (n=2181) | 834 (38.2%) | 1347 (61.8%) |  | **<0.001** |
| **Viral load (log10)** |  |  | **<0.001** |  |
| ≤ 4.0 (n=598) | 201 (33.6%) | 397 (66.4%) |  | **<0.001** |
| 4.1-5.0 (n=601) | 265 (44.1%) | 336 (55.9%) |  | **<0.001** |
| ≥ 5.1 (n=537) | 342 (63.7%) | 195 (36.3%) |  | **<0.001** |
| **ADR** |  |  | **0.010** |  |
| Yes (n=240) | 296 (61.3%) | 187 (38.7%) |  | **<0.001** |
| No (n=483) | 123 (51.2%) | 117 (48.8%) |  | 0.456 |
| **Subtype G** |  |  |  |  |
| **Sex** |  |  | 0.460 |  |
| Male (n=142) | 78 (54.9%) | 64 (45.1%) |  | 0.099 |
| Female (n=140) | 83 (59.3%) | 57 (40.7%) |  | **0.002** |
| **Treatment Status** |  |  | **0.015** |  |
| ART-naive (n=73) | 39 (53.4%) | 34 (46.6%) |  | 0.075 |
|  | LP | NLP | p-value | p-value compared |
| ART-experienced (n=92) | 66 (71.7%) | 26 (28.3%) |  | <0.001 |
| **Recentness of infection** |  |  | **0.003** |  |
| Chronic (n=152) | 99 (65.1%) | 53 (34.9%) |  | <0.001 |
| Recent (n=130) | 62 (47.7%) | 68 (52.3%) |  | 0.458 |
| **Transmission Route** |  |  | **0.035** |  |
| Heterosexual (n=42) | 22 (52.4%) | 20 (47.6%) |  | 0.660 |
| MSM (n=4) | 0 | 4 (100%) |  | 0.516 |
| IDU (n=25) | 17 (68%) | 8 (32%) |  | **0.011** |

## Discussion

In the current work, the use of the genomic sequences from the EuResist database combined with genomic sequences collected from public databases provides a comprehensive sample to study and characterize HIV-1 transmission clusters in Europe. On the one hand, HIV-1 transmission investigations are possible through reconstruction of transmission clusters. Information collected through such studies in a large scale can be highly useful for public health purposes, to fine-grain transmission patterns with higher resolution compared to classical epidemiology.

We reconstructed transmission clusters of HIV-1 in Europe with the specific objective of understanding the role of LP on transmission of infection. Specifically, we aimed to understand HIV-1 transmission clusters and determinants associated to transmission in clusters, taking into account the independent pandemics of the most prevalent subtypes in our population, A, B and C and to understand clustering patterns of LP and NLP in each subtype.

In our population the majority of patients were from subtype B, males, with MSM route of transmission and the region of origin of was Western Europe. These results are in accordance with a previous study conducted in Europe to analyse the distribution of subtypes (16).

We decided to study the transmission patterns of HIV-1 mainly according to subtypes, since there has been some discussion regarding the biological differences between them and mostly because of the higher prevalence of subtype B among Western Europe individuals (17). There were more patients outside TCs (78·4%) compared to those inside TCs (21·6%), in agreement with our study population based on sequences isolated at the first resistance test (18).

For subtype B, it was expected that one of the factors identified as associated with being inside a cluster was indeed the MSM transmission route (19). For subtype A, we also found MSM transmission route as a factor associated with being inside clusters. The fact that the MSM route of transmission is being associated with clustering in other non-B subtypes is in accordance with some studies that report an increase of non-B subtypes associated with MSM route (19–21). Nevertheless, we expected an association of IDU route of transmission and subtype A since both subtype and route of transmission are highly prevalent in Eastern Europe where this type of transmission route is also prevalent (22). On the other hand, in subtype G, being inside a cluster was associated with heterosexual transmission, as expected (23,24). We also found that, for subtype B, the age at resistance test was associated with being in cluster, with a higher probability among individuals with younger age. This is in accordance with some other studies (18,25). Migration status was also associated with being in clusters: migrants infected with subtype B were mainly in a cluster, and migrants infected with subtype G were less likely to be in a cluster. These results were in accordance with a recent study focusing on migration and HIV-1 in Portugal (26). These results could be explained by the region of origin of migrants. Subtype B had higher prevalence of migrants from Brazil, and Brazil has a concentrated HIV epidemic among MSM population (27,28).

Regarding the potential association between transmission clusters and late presentation, we found that both LP and NLP were mainly outside clusters. As for the differences between the populations of LP and NLP inside clusters, these patterns were consistent between subtypes A and B: concerning sex, there were more NLP males inside cluster; concerning age, there were older LP, with a higher and growing proportion as age increases. Subtype G had the most different patterns of all. Furthermore, for subtypes B and G individuals, there were more treated patients among LP than among NLP inside clusters. As for transmission route, for subtype G, we found more LP with an IDU transmission route inside clusters. Finally, for subtype B, it was interesting to observe that LP located inside clusters had higher viral loads than NLP. These results could not be compared by LP and NLP populations, nevertheless our results are overall in accordance with some studies (18,26,28).

Finally, we found that LP were more frequently present in small clusters or outside clusters compared to NLP which can indicate a limited role of this population on HIV-1 transmission, given the less frequent presence of these patients in TCs. However higher viral loads were observed in LP located inside clusters that can indicate higher transmissibility of infection within individuals from the TCs.

Finally, there is still scarce to none information regarding transmission clusters and late presentation. We studied here the association of transmission clusters according to subtypes in LP and NLP, and our results showed that the patterns of LP vs NLP in TCs presented similar characteristics in subtypes A and B, but not in subtype Gdominated by LP.

### Limitations

In our study we did not used the time and place of the most recent common ancester (tMRCA), instead we used a total number of sequences from a specific region. This methodology can cause some sampling bias since sequences can artificially be in cluster due to their shared region of origin.

## Conclusion

In conclusion, our study presented an updated description of the socio-demographic and clinical characteristics of HIV-1 infected individuals followed in Europe according to subtype. Our study also highlights the patterns of transmission clusters in LP vs NLP populations selected in the european dataset of EuResist. We conclude that late presentation could have a limited role on HIV-1 transmission. However further investigation should be considered to exclude LP classification bias, and to better estimate the time of infection based on phylogenetic trees reconstruction and molecular clock analysis.

## Data Availability

The original contributions presented in the study are included in the article/Supplementary Material, further inquiries can be directed to the corresponding author/s. The use of the EuResist database is available upon request through a study proposal submission.

https://www.euresist.org/eidb

## Author Contributions

Conceptualization, M.N.S.M., M.P. and A.A.; Methodology, M.N.S.M., M.P., V.P., M.d.R.O.M. and A.A.; Software, M.N.S.M., V.P., S.G.S.; Validation, M.N.S.M., M.P., F.I. and A.A.; Formal Analysis, M.N.S.M., V.P., S.G.S., M.P. and A.A.; Investigation, M.N.S.M., M.P., V.P., and M.d.R.O.M.; Resources, C.S.-D., R.P., R.K., M.B. (Marina Bobkova),M.B (Michael Böhm), M.Z., P.G and F.I.; Data Curation, C.S.-D., R.P., R.K., M.B. (Marina Bobkova),M.B (Michael Böhm), M.Z., P.G and F.I.; Writing—Original Draft Preparation, M.N.S.M., M.P. and A.A.; Writing— Review and Editing, M.N.S.M., M.P., C.S.-D., F.I. and A.A.; Visualization, M.N.S.M., M.P., V.P., M.d.R.O.M. and A.A.; Supervision, A.A.; Project Administration, A.A.; Funding Acquisition, A.A.

---

**Table Supplementary 1.** Patients socio-demographic and clinical characteristics

| Patient Characteristics | Total | Subtype A | Subtype B | Subtype G | p-value |
| --- | --- | --- | --- | --- | --- |
| **Total** | 38531 (100) | 2276 (5.9) | 32652 (84.7) | 3603 (9.4) |  |
| **Sex, n (%)** | 36699 (95.2) | 2011 (5.5) | 31200 (85.0) | 3488 (9.5) | <0.001 |
| Male | 27715 (75.5) | 1156 (4.2) | 24570 (88.6) | 1989 (7.2) |  |
| Female | 8984 (24.5) | 855 (9.5) | 6630 (73.8) | 1499 (16.7) |  |
| **Median age at resistance teste in years IQR, n (%)** | 18421 (47.8) | 631 (3.4) | 17149 (93.1) | 641 (3.5) | <0.001 |
|  | 40.0 (34.0-47.0) | 37.0 (30.0-45.0) | 41.0 (35.0-47.0) | 34.0 (28.0-41.0) |  |
| ≤ 18 | 233 (1.3) | 32 (13.7) | 143 (61.4) | 58 (24.9) | <0.001 |
| 19-30 | 2529 (13.7) | 150 (5.9) | 2209 (87.4) | 170 (6.7) |  |
| 31-55 | 14199 (77.1) | 390 (2.7) | 13430 (94.6) | 379 (2.7) |  |
| ≥ 56 | 1460 (7.9) | 59 (4.0) | 1367 (93.6) | 34 (2.4) |  |
| **Transmission Route, n (%)** | 18140 (47.1) | 1230 (6.8) | 16129 (88.9) | 781 (4.3) | <0.001 |
| Heterosexual | 5568 (30.7) | 532 (9.6) | 4646 (83.4) | 390 (7.0) |  |
| MSM | 6692 (36.9) | 97 (1.4) | 6566 (98.1) | 29 (0.4) |  |
| IDU | 4883 (26.9) | 433 (8.9) | 4269 (87.4) | 181 (3.7) |  |
| Other | 997 (5.5) | 168 (16.9) | 648 (65.0) | 181 (18.2) |  |
| **Region of origin, n (%)** | 23647 (61.4) | 1277 (5.4) | 20595 (87.1) | 1775 (7.5) | <0.001 |
| Western Europe | 20440 (86.4) | 482 (2.4) | 18836 (92.2) | 1122 (5.5) |  |
| Eastern Europe | 933 (3.9) | 468 (50.2) | 365 (39.1) | 100 (10.7) |  |
| Africa | 1162 (4.9) | 294 (25.3) | 343 (29.5) | 525 (45.2) |  |
| South America | 787 (3.3) | 11 (1.4) | 754 (95.8) | 22 (2.8) |  |
| Other | 325 (1.4) | 22 (6.8) | 297 (91.4) | 6 (1.8) |  |
| **Migration Status** | 16101 (41.8) | 574 (3.6) | 14629 (90.9) | 898 (5.6) | <0.001 |
| Migrant | 2543 (15.8) | 351 (13.8) | 1771 (69.6) | 421 (16.6) |  |
| Native | 13558 (84.2) | 223 (1.6) | 12858 (94.8) | 477 (3.6) |  |
| **Clusters** | 38531 (100) | 2276 (5.9) | 32652 (84.7) | 3603 (9.4) | 0.006 |
| In Clusters | 8335 (21.6) | 433 (5.2) | 7136 (85.6) | 766 (9.2) |  |
| Out-of-Clusters | 30196 (78.4) | 1843 (6.1) | 25516 (84.5) | 2837 (9.4) |  |
| **Treatment Status** | 21687 (56.3) | 709 (3.3) | 19387 (89.4) | 1591 (7.3) | <0.001 |
| ART-naïve | 8887 (41.0) | 443 (5.0) | 8046 (90.5) | 398 (4.5) |  |
| ART-experienced | 12800 (59.0) | 266 (2.1) | 11341 (88.6) | 1193 (9.3) |  |
| **Recentness of infection** | 38531 (100) | 2276 (5.9) | 32652 (84.7) | 3603 (9.4) | <0.001 |
| Recent | 15571 (40.4) | 929 (6.0) | 13322 (85.6) | 1320 (8.5) |  |
| Chronic | 22960 (59.6) | 1347 (5.9) | 19330 (84.2) | 2283 (9.9) |  |
| **TDR** | 7727 (20.1) | 393 (5.1) | 7035 (91.0) | 299 (3.9) | <0.001 |
| Yes | 967 (12.5) | 17 (1.8) | 918 (94.9) | 32 (3.3) |  |
| No | 6760 (87.5) | 376 (5.6) | 6117 (90.5) | 267 (3.9) |  |
| **ADR** | 6184 (16.0) | 133 (2.2) | 5320 (86.0) | 731 (11.8) | <0.001 |
| Yes | 4542 (73.4) | 73 (1.6) | 3958 (87.1) | 511 (11.3) |  |
| No | 1642 (26.6) | 60 (3.7) | 1362 (82.9) | 220 (13.4) |  |
| **Median CD4 count at diagnosis (cells/mL) IQR, n (%)** | 24321 (63.1) | 1011 (4.2) | 21583 (88.7) | 1727 (7.1) | <0.001 |
|  | 341.0 (170.0-540.0) | 328.0 (174.0-510.0) | 349.0 (172.0-547.0) | 273.0 (139.0-445.0) |  |
| LP | 12501 (51.4) | 545 (4.4) | 10875 (87.0) | 1081 (8.6) | <0.001 |
| NLP | 11820 (48.6) | 466 (3.9) | 10708 (90.6) | 646 (5.5) |  |
| **Viral Load at diagnosis (log<sub>10</sub> copies/mL) IQR, n (%)** | 15670 (40.7) | 614 (3.9) | 13105 (83.6) | 1951 (12.5) | <0.001 |
|  | 4.3 (3.4-5.0) | 4.3 (3.4-5.1) | 4.3 (3.3-5.0) | 4.5 (3.7-5.1) |  |
| ≤ 4.0 | 6210 (39.6) | 246 (4.0) | 5315 (85.6) | 649 (10.5) | <0.001 |
| 4.1-5.0 | 5361 (34.2) | 197 (3.7) | 4436 (82.7) | 728 (13.6) |  |
| ≥ 5.1 | 4099 (26.2) | 171 (4.2) | 3354 (81.8) | 574 (14.0) |  |

**Table Supplementary 2.** Unadjusted analysis for determinants associated with belonging to a transmission cluster according to Subtype A, B and G

|  |  | Subtype A |  | Subtype B |  | Subtype G |  |
| --- | --- | --- | --- | --- | --- | --- | --- |
| In clusters/Out-of-clusters |  | Unadjusted Model |  | Unadjusted Model |  | Unadjusted Model |  |
|  |  | uOR (95%CI) | p-value | uOR (95%CI) | p-value | uOR (95% CI) | p-value |
| Sex | Female | Ref | Ref | Ref | Ref | Ref | Ref |
|  | Male | 1.84 (1.46-2.32) | <0.001 | 1.49 (1.38-1.59) | <0.001 | 0.81 (0.69-0.96) | 0.012 |
| Age at resistance test | <18 | 0.56 (0.16-1.90) | 0.353 | 0.72 (0.45-1.17) | 0.187 | 1.85 (0.35-9.71) | 0.469 |
|  | 19-30 | 1.19 (0.57-2.49) | 0.641 | 1.72 (1.47-2.03) | <0.001 | 2.89 (0.65-12.80) | 0.162 |
|  | 31-55 | 0.76 (0.38-1.50) | 0.423 | 0.91 (0.80-1.06) | 0.223 | 2.16 (0.50-9.30) | 0.303 |
|  | >56 | Ref | Ref | Ref | Ref | Ref | Ref |
| Transmission Route | Heterosexual | Ref | Ref | Ref | Ref | Ref | Ref |
|  | MSM | 3.46 (2.12-5.65) | <0.001 | 1.84 (1.67-2.02) | <0.001 | 1.51 (0.62-3.67) | 0.367 |
|  | IDU | 0.76 (0.51-1.13) | 0.178 | 0.60 (0.53-0.68) | <0.001 | 1.52 (0.99-2.34) | 0.055 |
|  | Other | 1.06 (0.64-1.77) | 0.810 | 0.76 (0.60-0.97) | 0.025 | 0.76 (0.46-1.25) | 0.276 |
| Region of Origin | Western Europe | Ref | Ref | Ref | Ref | Ref | Ref |
|  | Eastern Europe | 0.63 (0.46-0.87) | 0.005 | 1.58 (1.26-1.99) | <0.001 | 0.67 (0.40-1.14) | 0.141 |
|  | Africa | 0.55 (0.37-0.80) | 0.002 | 1.11 (0.86-1.43) | 0.424 | 0.57 (0.43-0.75) | <0.001 |
|  | South America | 1.20 (0.31-4.59) | 0.793 | 0.94 (0.78-1.12) | 0.478 | 2.12 (0.90-5.02) | 0.087 |
|  | Other | 1.20 (0.46-3.13) | 0.714 | 1.02 (0.77-1.34) | 0.917 | 0.61 (0.07-5.27) | 0.656 |
| Migration Status | Migrant | Ref | Ref | Ref | Ref | Ref | Ref |
|  | Native | 1.48 (0.97-2.27) | 0.070 | 0.72 (0.64-0.81) | <0.001 | 1.57 (1.09-2.26) | 0.015 |
| Recentness of Infection | Chronic | Ref | Ref | Ref | Ref | Ref | Ref |
|  | Recent | 2.70 (2.18-3.35) | <0.001 | 2.22 (2.10-2.34) | <0.001 | 2.31 (1.96-2.72) | <0.001 |
| ADR | Yes | Ref | Ref | Ref | Ref | Ref | Ref |
|  | No | 2.01 (0.87-4.64) | 0.102 | 1.54 (1.30-1.82) | <0.001 | 2.35 (1.47-3.74) | <0.001 |
| LP/NLP | LP | Ref | Ref | Ref | Ref | 1 | 1 |
|  | NLP | 1.61 (1.16-2.23) | 0.004 | 1.15 (1.08-1.23) | <0.001 | 1.32 (1.02-1.71) | 0.037 |
| Viral load groups | <4.0 | Ref | Ref | Ref | Ref | 1 | 1 |
|  | 4.1-5.0 | 1.15 (0.67-1.98) | 0.603 | 1.27 (1.14-1.42) | <0.001 | 1.16 (0.88-1.53) | 0.297 |
|  | >5.1 | 1.66 (0.98-2.82) | 0.060 | 1.55 (1.38-1.73) | <0.001 | 1.24 (0.93-1.66) | 0.144 |

